# Healthcare resource utilisation and costs of hospitalisation and primary care among adults with COVID-19 in England: a population-based cohort study

**DOI:** 10.1101/2023.05.10.23289557

**Authors:** Jingyan Yang, Kathleen M. Andersen, Kiran K. Rai, Theo Tritton, Tendai Mugwagwa, Maya Reimbaeva, Carmen Tsang, Leah J. McGrath, Poppy Payne, Bethany Backhouse, Diana Mendes, Rebecca Butfield, Kevin Naicker, Mary Araghi, Robert Wood, Jennifer L. Nguyen

**Author notes:** **Corresponding Author:** Jingyan Yang, DrPH MHS, Global Value and Access, Pfizer Inc., 66 Hudson Blvd E, New York, NY 10001, USA.

## Abstract

**Objectives:** To quantify healthcare resource utilisation (HCRU) and costs to the National Health Service (NHS) associated with acute COVID-19 in adults in England.

**Design:** Population-based retrospective cohort study, using Clinical Practice Research Datalink (CPRD) Aurum primary care electronic medical records linked when available to Hospital Episode Statistics (HES) secondary care administrative data.

**Setting:** Patients registered to primary care practices in England.

**Population:** 1,706,368 adults with a positive SARS-CoV-2 PCR or antigen test from August 2020 to January 2022 were included; 13,105 within the hospitalised cohort indexed between August 2020 and March 2021, and 1,693,263 within the primary care cohort indexed between August 2020 and January 2022.

**Main outcome measures:** Primary and secondary care HCRU and associated costs during the acute phase of COVID-19 (≤4 weeks following positive test), stratified by age group, risk of severe COVID-19 and immunocompromised status.

**Results:** Among the hospitalised cohort, average total length of stay, as well as in critical care wards, was longer in older adults. Median healthcare cost per hospitalisation was higher in those aged 75 – 84 (£8,942) and ≧85 years (£8,835) than in those aged <50 years (£7,703). Whilst few (6.0%) patients in critical care required mechanical ventilation, its use was higher in older adults (50 – 74 years: 8.3%; <50 years: 4.3%). HCRU and associated costs were often greater in those at higher risk of severe COVID-19 when compared to the overall cohort, although minimal differences in HCRU were found across the three different high-risk definitions implemented. Among the primary care cohort, GP or nurse consultations were more frequent among older adults and the immunocompromised.

**Conclusions:** COVID-19 related hospitalisations in older adults, particularly critical care admissions, were the primary drivers of high resource use of COVID-19 in England. These findings may inform health policy decisions and resource allocation in the prevention and management of COVID-19.

## INTRODUCTION

Coronavirus disease 2019 (COVID-19) is a highly infectious respiratory illness caused by the severe acute respiratory syndrome-coronavirus-2 virus (SARS-CoV-2). Globally, ^∼^670 million cases and ^∼^6.9 million deaths related to COVID-19 have been recorded as of 9^th^ March 2023.^1^ Within England, as of 1^st^ March 2023 there have been approximately 20.5 million cases, 982,000 hospitalisations and 186,000 associated deaths.^2^ The clinical presentation of COVID-19 ranges from asymptomatic to critical illness, where mild or uncomplicated illness is commonly managed in primary care and severe COVID-19 managed in the hospital setting.^3 4^ Whilst the majority experience few symptoms or mild to moderate COVID-19, some patients require medical intervention, including respiratory support and intensive care admission.^5^ The risk of worse outcomes, e.g. hospitalisation and death, is greater for older adults, smokers, those who are obese, have a compromised immune system, and/or have certain comorbidities such as hypertension and lung disease.^6 7^ As such, the Joint Committee on Vaccination and Immunisation (JCVI) recommended the prioritisation of COVID-19 vaccination in specific groups (based on age, those clinically extremely vulnerable, underlying health conditions, pregnancy and working in health and social care).^8^

Whilst the vaccination roll-out has substantially reduced COVID-19 associated morbidity and mortality, COVID-19 remains a significant burden on the UK healthcare system. According to a report published by the UK’s Department of Health and Social Care (DHSC) assessing the impacts during the Omicron wave in England, COVID-19 has led to longer waits for elective and emergency visits in the secondary care setting, and across the pandemic COVID-19 has reduced or delayed appointments and referrals in primary care, potentially resulting in a worsened state of health for some primary care patients.^9^

In addition to the health impact of COVID-19, several studies in the United States (US) have quantified the economic burden of COVID-19 related hospitalisations,^10-12^ and have reported higher costs among those with health complications. Studies quantifying the economic burden of COVID-19 in the UK are scarce; of the 37 studies identified within a systematic review quantifying the economic impact of COVID-19, five focused on UK-based data, of which two assessed direct healthcare burden attributed to patient care, three assessed the macroeconomic impact of the epidemic and its associated policies and two assessed the costs associated with COVID-19.^13^ Only two studies reported use of electronic health records, with no studies reporting the use of general practitioner (GP) appointments data. Keogh-Brown *et al*. report that the impact of COVID-19 on the UK economy was approximately a loss of £40 billion in 2020.^14^ However, much of these costs were related to reduced labour (^∼^£39 billion), and whilst overall hospital and intensive care costs were estimated (^∼^£1 billion), these focused on the health-related economic impact on to the UK economy, rather than individual-level costs to the NHS to manage patients with COVID-19. Furthermore, to our knowledge, no studies have reported direct medical costs within the primary care setting. By addressing these data gaps within the literature, we will provide valuable evidence to support health policy decisions on public health interventions and healthcare resource allocation in the prevention and management of COVID-19.

### Aims and objectives

This study aimed to quantify healthcare resource utilisation (HCRU) and costs associated with COVID-19 in adults in England, by age and according to risk of severe COVID-19 and immunocompromised status, separately for those with and without hospitalisation records, using UK primary care data, linked to secondary care data when available.

## METHODS

### Study design and setting

We conducted a population-based retrospective cohort study using data obtained from the Clinical Practice Research Datalink (CPRD) Aurum primary care database^15^ and, when available, linked secondary care data (Hospital Episode Statistics Admitted Patient Care dataset, HES APC).^16^ The May 2022 release of CPRD Aurum was used; the latest data from CPRD Aurum covers the period January 1995 to April 2022,^17 18^ while HES APC covers April 1997 to March 2021.^16^ The study design and methods have been described elsewhere.^19^ A study design schematic is provided in Supplementary eFigure 1.

Two distinct patient cohorts were created to describe the economic burden of the acute phase of COVID-19 among adults in the UK:

1. **Hospitalised cohort:** Patients who had a positive SARS-CoV-2 polymerase chain reaction (PCR) or antigen test, or a recorded clinical diagnosis of COVID-19, in their GP record between 1 August 2020 and 31 March 2021 and had a COVID-19 related hospitalisation within 84 days after their positive test result. The index period start date was chosen to align with when it became mandatory for National Health Service (NHS) Test and Trace to report positive PCR test results to the patient’s general practitioner (GP) practice, from 20^th^ July 2020 onwards;^20^ the end date was determined by the end of data availability within HES APC. Patients in this cohort may have also received COVID-19 care outside of the hospital setting e.g. primary care consultations.
2. **Primary care cohort:** Patients who had a positive SARS-CoV-2 PCR or antigen test, or a recorded clinical diagnosis of COVID-19, in their GP record. For persons diagnosed between 1 August 2020 and 31 March 2021, they were included in this cohort if they did not have a record for a COVID-19 related hospitalisation within 84 days of their positive test result. All persons diagnosed with COVID-19 on or after 1 April 2021 were included in this cohort, as this was a period of time for which CPRD did not have hospitalisation data available. The index period end date, January 2022, was determined by the overall cohort design.^19^

### Population

Patients aged ≥18 years were included in this study. Further details on the eligibility criteria are cited elsewhere.^19^ In brief, this study included patients that had: 1) a confirmed COVID-19 episode recorded in CPRD Aurum, where the first date of COVID-19 diagnosis (i.e., index date) was observed in the index period, 2) a minimum registration period of 12 months at their current GP practice prior to the index date, 3) data considered of acceptable research quality as defined by CPRD,^21^ and 4) eligible for linkage to HES. Patients were excluded if they had a record for a COVID-19 related hospitalisation or death prior to their GP-recorded date of COVID-19 diagnosis. COVID-19 episodes starting prior to August 2020, from which point capture of COVID-19 test results within GP patient records was considered nearly complete, were not included in the study.

### Demographic and clinical characteristics

Sociodemographic characteristics at index included: age, sex, region of GP practice, ethnicity (using patient history), social deprivation (measured using the Index of Multiple Deprivation (IMD) 2019 score), smoking status (using patient history) and body mass index (BMI) within two years of the index date. Clinical characteristics included Quan-Charlson Comorbidity Index (CCI) 2005^22^ within two years of index, and vaccination status according to immunocompromised status at index. Vaccination status at index for immunocompetent patients was defined according to whether they had received 0 doses (unvaccinated), 1 primary dose, 2 primary doses, or any booster dose. For immunocompromised patients, vaccination status was defined according to whether they had received 0 doses (unvaccinated), 1 primary dose, 2 primary doses, 3 primary doses, or any booster dose. A patient was considered vaccinated starting from 14 days after dose receipt, and doses were required to be separated by at least 21 days. Disease severity among the hospitalised cohort was assessed using the Ordinal Scale for Clinical Improvement within WHO’s COVID-19 Therapeutic Trial Synopsis,^23^ based on the highest level of care received during the hospitalisation following mutually exclusive categories: 1) hospitalised, no oxygen therapy; 2) oxygen by mask or nasal prongs; 3) noninvasive ventilation or high-flow oxygen; 4) intubation and mechanical ventilation and 5) ventilation and additional organ support. Further details on definitions, and operationalisation of code lists are described elsewhere.^19^

### Outcomes and follow-up

#### Healthcare resource utilisation (HCRU)

All COVID-19 related HCRU and associated costs to the NHS in the four weeks including and following the index date were calculated and reported for the following elements of HCRU:

##### Medication use

Medications that were prescribed within primary care were considered to be COVID-19 related when prescribed on the same day as a COVID-19 diagnosis (see Supplementary eTable 1).^24 25^

**Table 1:** Demographic and clinical characteristics of the study population at baseline.

| Characteristics | Hospitalised cohort<br>(n=13,105)<br>n (%) | Primary care cohort (n=1,693,263)<br>n (%) |
| --- | --- | --- |
| **Age, years** |  |  |
| Mean (SD) | 60.7 (16.0) | 42.3 (15.8) |
| 18-49 | 3,127 (23.9%) | 1,161,843 (68.6%) |
| 50-64 | 4,844 (37.0%) | 379,528 (22.4%) |
| 65-74 | 2,386 (18.2%) | 92,573 (5.5%) |
| 75-84 | 1,690 (12.9%) | 40,481 (2.4%) |
| ≥85 | 1,058 (8.1%) | 18,838 (1.1%) |
| **Sex** |  |  |
| Male | 7,504 (57.3%) | 764,687 (45.2%) |
| Female | 5,601 (42.7%) | 928,546 (54.8%) |
| Unknown | 0 | 30 (<0.1%) |
| **GP practice region** |  |  |
| North East | 465 (3.6%) | 65,755 (3.9%) |
| North West | 2,872 (21.9%) | 363,770 (21.5%) |
| Yorkshire and The Humber | 339 (2.6%) | 53,777 (3.2%) |
| East Midlands | 213 (1.6%) | 38,663 (2.3%) |
| West Midlands | 2,057 (15.7%) | 278,170 (16.4%) |
| East of England | 413 (3.2%) | 66,446 (3.9%) |
| London | 2,965 (22.6%) | 335,233 (19.8%) |
| South East | 2,670 (20.4%) | 317,898 (18.8%) |
| South West | 1,111 (8.5%) | 173,317 (10.2%) |
| Unknown | 0 | 234 (<0.1%) |
| **Ethnicity** |  |  |
| White | 8,769 (66.9%) | 1,106,974 (65.4%) |
| Black | 467 (3.6%) | 45,177 (2.7%) |
| Asian | 1,651 (12.6%) | 108,603 (6.4%) |
| Mixed | 108 (0.8%) | 17,757 (1.1%) |
| Other | 329 (2.5%) | 34,726 (2.1%) |
| Unknown | 1,781 (13.6%) | 380,026 (22.4%) |
| **Index of Multiple Deprivation (2019)** |  |  |
| Quintile 1 (least deprived) | 2,166 (16.5%) | 335,374 (19.8%) |
| Quintile 2 | 2,342 (17.9%) | 336,418 (19.9%) |
| Quintile 3 | 2,488 (19.0%) | 325,643 (19.2%) |
| Quintile 4 | 2,881 (22.0%) | 353,803 (20.1%) |
| Quintile 5 (most deprived) | 3,221 (24.6%) | 340,894 (20.1%) |
| Unknown | 7 (<0.1%) | 1,131 (0.1%) |
| **Smoking** |  |  |
| Current smoker | 1,690 (12.9%) | 293,016 (17.3%) |
| Ex-smoker | 3,810 (29.1%) | 283,693 (16.8%) |
| Never smoked | 4,196 (32.0%) | 532,804 (31.5%) |
| Unknown | 3,409 (26.0%) | 583,750 (34.5%) |
| **BMI** |  |  |
| Underweight | 94 (0.7%) | 13,657 (0.8%) |
| Normal | 1,154 (8.8%) | 201,500 (11.9%) |
| Overweight | 2,388 (18.2%) | 198,235 (11.7%) |
| Obese | 4,586 (35.0%) | 215,802 (12.7%) |
| Unknown | 4,883 (37.3%) | 1,064,069 (62.8%) |
| **Quan Charlson Comorbidity Index (2005)** |  |  |
| Mean (SD) | 1.0 (1.7) | 0.2 (0.7) |
| 0 | 7,078 (54.0%) | 1,462,791 (86.4%) |
| 1-2 | 4,232 (32.3%) | 197,557 (11.7%) |
| 3-4 | 1,160 (8.9%) | 24,240 (1.4%) |
| ≥5 | 635 (4.9%) | 8,675 (0.5%) |
| **Immunocompromised status** |  |  |
| Immunocompetent | 12,924 (98.6%) | 1,629,716 (96.2%) |
| Immunocompromised | 181 (1.4%) | 63,547 (3.8%) |
| **Vaccination status: immunocompetent** |  |  |
| Unvaccinated | 12,684 (98.1%) | 660,610 (40.5%) |
| 1 dose | 240 (1.9%) | 125,129 (7.7%) |
| 2 doses | 0 | 593,557 (36.4%) |
| First booster dose | 0 | 250,420 (15.4%) |
| **Vaccination status: immunocompromised** |  |  |
| Unvaccinated | 102 (56.4%) | 1,154 (1.8%) |
| 1 dose | 79 (43.7%) | 5,314 (8.4%) |
| 2 doses | 0 | 33,731 (53.1%) |
| 3 doses | 0 | 23,082 (36.3%) |
| First booster dose | 0 | 266 (0.4%) |
| **Risk of severe COVID-19** |  |  |
| McInnes Advisory Group | 4,323 (33.0%) | 250,329 (14.8%) |
| PANORAMIC | 11,011 (84.0%) | 691,593 (40.8%) |
| Green Book | 5,341 (40.8%) | 212,556 (12.6%) |
| **COVID-19 severity<sup>a</sup>** |  |  |
| No oxygen therapy | 9,606 (73.3%) | - |
| Low-flow oxygen | 793 (6.1%) | - |
| Non-invasive ventilation or high-flow oxygen | 1,859 (14.2%) | - |
| Intubation/mechanical ventilation | 822 (6.3%) | - |
| Ventilation and additional organ support | 25 (0.2%) | - |

<sup>a</sup> Highest level of severity experienced during the hospitalisation.

##### Primary care consultations

GP or nurse consultations with a diagnostic code of COVID-19 were reported separately for face-to-face (F2F) and telephone consultations. This was defined as a maximum of one visit of each format per person per day, and any additional visits were considered as data capture errors.

##### Hospitalisations

Hospital admissions with a COVID-19 primary diagnosis were assessed within the hospitalised cohort. Additionally, the mean (standard deviation) and median (interquartile range) length of stay (LoS) per admission was assessed and reported separately for time spent in hospital from admission to discharge as well as time spent in high dependency/intensive care units (HDUs/ICUs). Whether mechanical ventilation treatment was received was also reported. In the event of multiple hospitalisations (or for critical care, HDU/ICU stays) during the acute COVID-19 phase, the average LoS per person, rather than the cumulative total, was used.

##### Direct healthcare costs

Costs were described for patients with one or more event of a given type only, i.e. resource users, and persons without utilisation were not included in the distributions of costs presented. The estimation of costs associated with hospitalisations was based on the National Schedule of NHS Costs (2020/2021) which report costs of admitted patient care by Healthcare Resource Group (HRG) in England.^26^ In order to estimate the cost per hospitalisation, all finished consultant episodes (FCEs: the time a patient spends in the care of one consultant within their hospitalisation) within one admission were accounted to derive the total spell (hospitalisation) cost.^27^ In the event that a given person had multiple hospitalisations during the acute phase, cost per day estimates were obtained by dividing total hospitalisation costs by total LoS per-patient.

Primary care consultations (including GP or nurse visits) were costed using information compiled and provided by the Personal Social Services Research Unit (PSSRU).^28^ The direct healthcare cost for each prescription written in primary care was calculated via the application of cost per unit from the NHS Drug Tariff.^29^

### Statistical analysis

This descriptive study included all patients who met the study eligibility criteria. This study did not involve hypothesis testing; therefore, formal sample size calculations were not performed. Means and standard deviation (SD), or median and lower and upper quartile (Q1, Q3) were calculated for numeric variables, with frequency counts and percentages presented for categorical variables. Per the design of the study, all results were presented separately for the hospitalised cohort and primary care cohorts.

### Stratifying variables

All outcomes were evaluated by age, high risk status, and immunocompromised status, as previous studies have shown that healthcare utilisation can differ by age and clinical status.^30 31^ Age group categories were based on the COVID-19 vaccination rollout strategy in the UK: 18 – 49; 50 – 64; 65 – 74; 75 – 84; and 85+. Three separate definitions were used to define persons at greater risk of severe COVID-19: 1) the McInnes Advisory Group highest risk group (a list of conditions to identify persons at the very highest risk of COVID hospitalisation and death, as defined by an advisory group chaired by Prof Iain McInnes and supported by the NHS England RAPID-C19 team (the McInnes Advisory Group)),^32^ 2) eligibility for the PANORAMIC study (a randomised trial of antiviral therapeutic agents including patients who were deemed at a higher risk of hospitalisation and death (PANORAMIC))^33^ and 3) the UK Health Security Agency (UKHSA) clinical groups, outlined in COVID-19: the Green Book, chapter 14a (JCVI’s COVID-19 vaccination prioritisation criteria) (the Green Book).^8^ The code lists for each high risk definition were developed using reproducible search terms with multiple reviewers, have been previously described and published.^19^ For immune system status, patients were classified as immunocompromised at the time of receipt of first COVID-19 vaccine dose if they had one or more codes meeting Davidson *et al*.’s definition of immunocompromised status.^34^ All analyses were conducted in SAS version 9.4 (SAS Institute, Cary, North Carolina).

### Patient and Public Involvement

There was no direct involvement of patients and public in this study. However, we aim to disseminate the findings through appropriate channels.

## RESULTS

### Patient sociodemographic and clinical characteristics

A total of 1,706,368 adult COVID-19 cases were included in this study. Of the 471,128 patients diagnosed between 1^st^ August 2020 and 31^st^ March 2021, 13,105 (2.8%) patients were included in the hospitalised cohort; 1,693,263 were included in the primary care cohort. **Table 1** summarises the baseline patient characteristics across the hospitalised and primary care cohorts.

Among the hospitalised cohort, the majority (n=9,978; 76.1%) of patients were aged ≥50 years (mean age [SD]: 60.7 [16] years), more than half (n=7,504; 57.3%) were male, 66.9% were of white ethnic origin, and 46.6% (n=6,102) lived in areas with greatest deprivation (IMD quintiles 4 and 5).

Of those with known smoking status, 56.7% (n=5,500) had a history of smoking. Over half (n=6,974; 53.2%) were overweight or obese according to BMI, and among those with a BMI record, this increased to 84.8%. Over half of patients (n=7,078; 54.0%) had a CCI score of 0. The majority (98.6%) of the hospitalised cohort were defined as immunocompetent in baseline, of whom 98.1% were unvaccinated at index (n=12,684; 98.1%).

The proportions of patients meeting the McInnes Advisory Group, PANORAMIC and the Green Book definitions, respectively were 33.0% (n=4,323), 84.0% (n=11,011) and 40.8% (n=5,341). Most (73.3%) patients did not receive oxygen therapy during their hospitalisation, and few patients (n=847; 6.5%) received intubation or ventilation support.

Sociodemographic characteristics of the primary care cohort differed numerically from the hospitalised cohort: the majority (n=1,161,843; 68.6%) of patients were aged <50 years (mean age [SD]: 42.3 [15.8] years), more than half (n=928,546; 54.8%) were female, and patients were relatively evenly spread across socioeconomic quintiles. However, the distribution across ethnicity groups was similar to the hospitalised cohort.

When considering the clinical characteristics, among those with known smoking status 52.0% had a smoking history, and 24.4% were overweight or obese (n=414,037). Most patients (n=1,462,791; 86.4%) had a CCI score of 0. Unlike the hospitalised cohort, vaccination status was more varied; among the 1,629,716 (96.2%) immunocompetent patients in the primary care cohort, 40.5% were unvaccinated.

Relatively fewer patients than in the hospitalised cohort were at risk of severe COVID-19 across all three definitions (McInnes Advisory Group: n=250,329, 14.8%; PANORAMIC: n=691,593, 40.8% and the Green Book: n=212,556, 12.6%).

### Healthcare resource utilisation (HCRU) and associated costs

#### Hospitalised cohort

The median total spell length of stay (LoS), including both general ward as well as critical care admission, was 6.0 days. The LoS was longer for older patients; median length of stay was 5.0 days for those aged 18 – 49 years, 6.0 days for age 50 – 64 years, and 8.0 days for age ≥65 years (**Table 2**). When stratified by risk of severe COVID-19, LoS was similar across definitions (median 7.0 days [Q1: 4.0, Q3: 12.0]). The median LoS was one day longer in immunocompromised patients (7.0 [4.0, 12.0]) compared to those immunocompetent (6.0 [4.0, 12.0]).

**Table 2:** HCRU in the four weeks after index stratified by age, risk of severe COVID-19 and immunocompromised status at baseline among the hospitalised cohort.

| HCRU |  | Age stratifications |  |  |  |  | Risk criteria |  |  | Immunocompromised status at baseline |  |
| --- | --- | --- | --- | --- | --- | --- | --- | --- | --- | --- | --- |
|  | All<br>(n=13,105) | 18-49<br>(n=3,127) | 50-64<br>(n=4,844) | 65-74<br>(n=2,386) | 75-84<br>(n=1,690) | ≥85<br>(n=1,058) | McInnes<br>Advisory<br>Group<br>(n=4,323) | PANORAMIC<br>(n=11,011) | Green Book<br>(n=5,341) | Immuno-<br>compromised<br>(n=181) | Immuno-<br>competent<br>(n=12,924) |
| **Length of hospital stay (in days)** |  |  |  |  |  |  |  |  |  |  |  |
| Mean (SD) | 9.2<br>(10.0) | 6.4<br>(8.1) | 9.0 (10.1) | 10.8 (10.9) | 11.2 (10.2) | 11.4 (9.7) | 9.9 (10.0) | 9.8 (10.2) | 10.0<br>(9.8) | 9.0<br>(6.7) | 9.2 (10.0) |
| Median (Q1, Q3) | 6.0<br>(4.0, 11.0) | 5.0<br>(2.0, 7.0) | 6.0<br>(4.0, 10.0) | 8.0<br>(5.0, 13.0) | 8.0<br>(5.0, 14.0) | 8.0<br>(5.0, 15.0) | 7.0<br>(4.0, 12.0) | 7.0<br>(4.0, 12.0) | 7.0<br>(4.0, 12.0) | 7.0<br>(4.5, 12.0) | 6.0<br>(4.0, 11.0) |
| **Critical care admission** |  |  |  |  |  |  |  |  |  |  |  |
| n (%) | 1,934 (14.8%) | 399 (12.8%) | 896 (18.5%) | 456 (19.1%) | 165 (9.8%) | 18<br>(1.7%) | 608 (14.1%) | 1,687 (15.3%) | 695 (13.0%) | 10 (5.5%) | 1,924 (14.9%) |
| **Length of stay in critical care** |  |  |  |  |  |  |  |  |  |  |  |
| Mean (SD) | 11.5 (11.2) | 9.7 (10.9) | 12.3 (11.8) | 12.3 (11.1) | 10.0<br>(8.9) | 5.3<br>(5.6) | 11.4 (10.9) | 11.7 (11.1) | 11.2 (10.6) | 9.0<br>(5.8) | 11.5 (11.3) |
| Median (Q1, Q3) | 8.0<br>(4.0, 15.0) | 6.0<br>(4.0, 11.0) | 8.5<br>(5.0, 16.0) | 9.0<br>(5.0, 15.0) | 8.0<br>(4.0, 14.0) | 4.0<br>(1.0, 7.0) | 8.0<br>(5.0, 15.0) | 8.0<br>(5.0, 15.0) | 8.0<br>(5.0, 14.0) | 8.5<br>(4.0, 13.0) | 8.0<br>(4.2, 15.0) |
| **Mechanical ventilation** |  |  |  |  |  |  |  |  |  |  |  |
| n (%) | 792<br>(6.0%) | 135<br>(4.3%) | 398<br>(8.2%) | 204<br>(8.6%) | 55<br>(3.3%) | 0 | 250<br>(5.8%) | 713<br>(6.5%) | 278<br>(5.2%) | <5 | 790<br>(6.1%) |
| **Mechanical ventilation days** |  |  |  |  |  |  |  |  |  |  |  |
| Mean (SD) | 1.2<br>(0.5) | 1.2<br>(0.4) | 1.2<br>(0.4) | 1.2<br>(0.5) | 1.1<br>(0.4) | 0 | 1.2<br>(0.4) | 1.2<br>(0.5) | 1.2<br>(0.4) | N/A* | 1.2<br>(0.5) |
| Median (Q1, Q3) | 1.0<br>(1.0, 1.0) | 1.0<br>(1.0, 1.0) | 1.0<br>(1.0, 1.0) | 1.0<br>(1.0, 1.0) | 1.0<br>(1.0, 1.0) | 0 | 1.0<br>(1.0, 1.0) | 1.0<br>(1.0, 1.0) | 1.0<br>(1.0, 1.0) | N/A* | 1.0<br>(1.0, 1.0) |

In the event of multiple hospitalisations (or critical care stays) during the acute COVID-19 phase, the average per person, rather than the cumulative total, was used.
\*Suppressed to comply with the reporting rules.

**Table 3:**
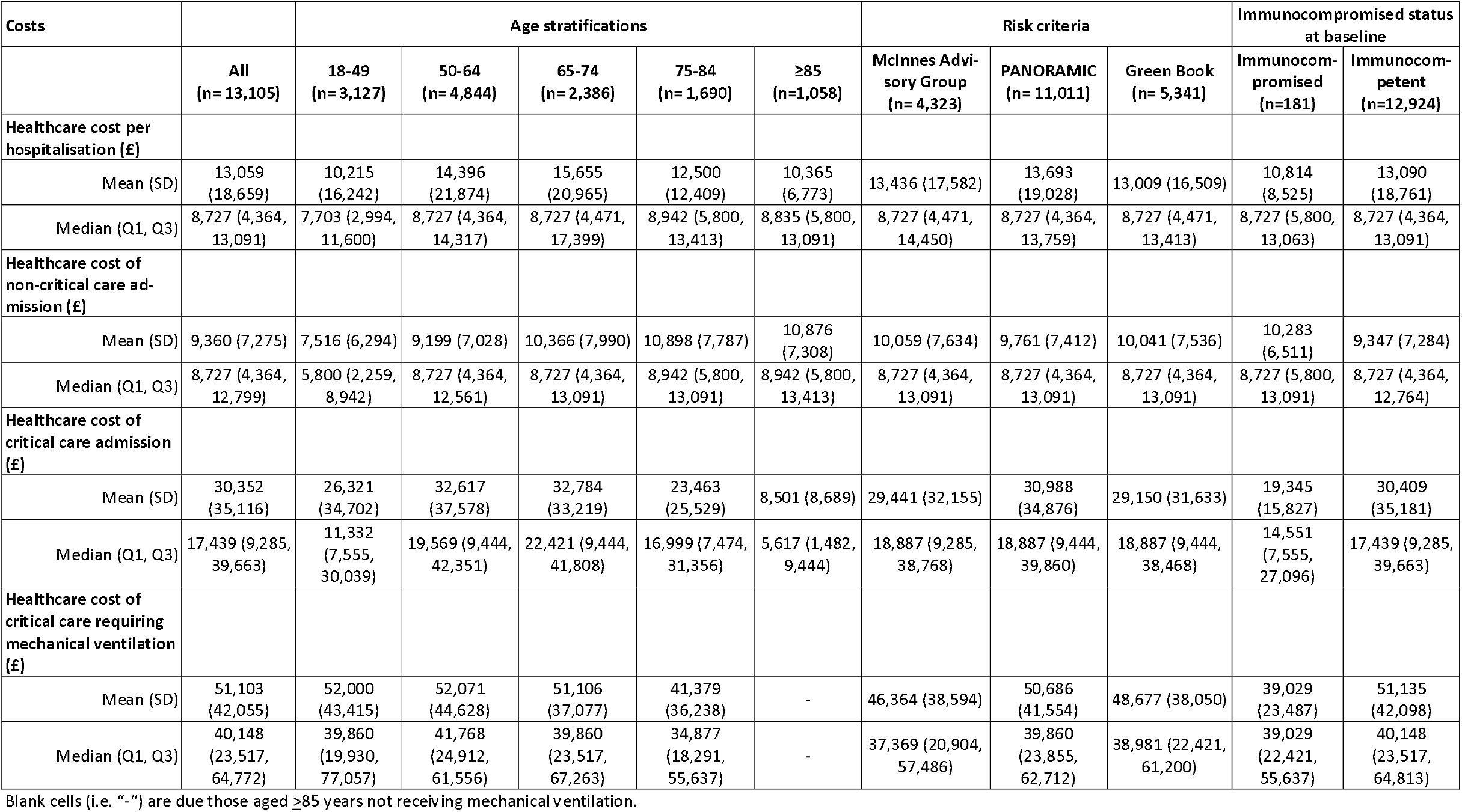
Costs in the four weeks after index stratified by age, risk of severe COVID-19 and immunocompromised status at baseline among the hospitalised cohort.

Of the 13,105 hospitalised patients, 1,934 (14.8%) were admitted to critical care. The median LoS in critical care was 8.0 days. The proportion requiring critical care, as well as LoS, was greatest among the aged 50-64 and 65-74 groups. Critical care LoS was similar for persons meeting each risk definitions as well as by immunocompromised status when compared to the overall cohort.

Median healthcare cost per hospitalisation was lower in those aged 18 – 49 years (£7,703) than in those aged 75 – 84 years (£8,942) and <u>≥</u>85 years (£8,835), and was similar across the risk definitions (£8,727) and immunocompromised status (£8,727 in the immunocompromised and immunocompetent groups). The median non-critical care costs followed similar patterns, whereas median observed critical care costs were higher among immunocompetent patients (£17,439) than among immunocompromised patients (£14,551). The mean number of FCEs per hospitalisation was 2.0, with FCEs ranging between 1-7 for immunocompromised patients and 1-16 patients for immunocompetent patients (data not shown in tables).

Median costs of critical care requiring mechanical ventilation (MV) was broadly similar across all stratifications. Overall, the proportion of hospitalised patients who received MV was low (n=792; 6.0%) and increased with age (4.3%, for ages 18 – 49, 8.2% for ages 50 – 64 and 8.6% for ages 65 – 74 years), but decreased after age 74. MV use varied slightly across the risk definitions, with the highest use in the PANORAMIC group (6.5%), and lowest in the Green Book group (5.2%). Among people who received MV, the median length of ventilation was 1.0 day; this did not differ across stratifications.

Telephone consultations with a GP or nurse (n=5,077; 38.7%) were more common than F2F consultations (n=2,489; 19.0%) (Supplementary materials: eTable 2). Telephone visits remained the main mode of consultation when stratified by age and risk definition, particularly for older adults, people at high risk of severe COVID-19 and immunocompromised patients. When assessing COVID-19 associated medication use, only 29 (0.2%) patients received a primary care prescription from their GP to manage or treat COVID-19 (Supplementary materials eTable 2).

#### Primary care cohort

The proportion of patients with ≥1 F2F GP or nurse consultation was higher in the older age groups (aged ≥85 years: 12.7%; aged 18 – 49 years: 3.4%;) (**Table 4**). Similar patterns were observed for ≥1 telephone consultations, however greater use was observed across all ages (aged 18 – 49 years: 6.1%; aged ≥85 years: 25.8%) when compared to F2F consultations.

**Table 4:** HCRU in the four weeks after index stratified by age, risk of severe COVID-19 and immunocompromised status at baseline among the primary care cohort.

| HCRU |  | Age stratifications |  |  |  |  | Risk criteria |  |  | Immunocompromised status at baseline |  |
| --- | --- | --- | --- | --- | --- | --- | --- | --- | --- | --- | --- |
|  | All<br>(n=1,693,263) | 18-49<br>(n=1,161,843) | 50-64<br>(n=379,528) | 65-74<br>(n=92,573) | 75-84<br>(n=40,481) | ≥85<br>(n=18,838) | McInnes Advisory Group<br>(n=250,329) | PANORAMIC<br>(n=691,593) | Green Book<br>(n=212,556) | Immunocompromised<br>(n=63,547) | Immunocompetent<br>(n=1,629,716) |
| **Any COVID-19 medication use:** n (%) | 253<br>(<1.0%) | 69<br>(<1.0%) | 63<br>(<1.0%) | 33<br>(<1.0%) | 39<br>(0.1%) | 49<br>(0.3%) | 158<br>(0.1%) | 214<br>(<1.0%) | 116 (0.1%) | 51<br>(0.1%) | 202<br>(<1.0%) |
| **Primary care consultations-F2F:** number with ≥ 1 visit (%) | 71,039<br>(4.2%) | 39,108 (3.4%) | 19,330 (5.1%) | 6,491 (7.0%) | 3,726 (9.2%) | 2,384 (12.7%) | 16,989 (6.8%) | 40,991 (5.9%) | 17,237<br>(8.1%) | 4,587<br>(7.2%) | 66,452 (4.1%) |
| **Primary care consultations-Telephone:** number with ≥ 1 call (%) | 137,148<br>(8.1%) | 70,675 (6.1%) | 40,311<br>(10.6%) | 13,434<br>(14.5%) | 7,860 (19.4%) | 4,868 (25.8%) | 35,935<br>(14.4%) | 86,002<br>(12.4%) | 36,852<br>(17.3%) | 9,375<br>(14.8%) | 127,773<br>(7.8%) |

Among COVID cases managed in the community, none had record of receipt of nirmatrelvir/ritonavir, remdesivir, casirivimab/imdevimab or baricitinib.

When assessing those at risk of severe COVID-19, we observed a similar proportion of patients with ≥1 F2F consultations across the three risk definitions (McInnes Advisory Group: 6.8%; PANORAMIC: 5.9% and the Green Book: 8.1%). However, the patterns of ≥1 telephone-based consultation slightly differed across the three risk definitions; with highest use noted for the Green Book criteria (17.3%) and lowest for the PANORAMIC criteria (12.4%). Of immunocompromised patients, 7.2% had at least one F2F consultation and 14.8% had at least one telephone consultation, in comparison to 4.1% and 7.8% for immunocompetent patients, respectively.

Among the primary care cohort, <1.0% (n=253) of patients received a primary care prescription for treatment of COVID-19.

The overall median costs were higher for F2F consultations compared to a telephone consultation among those with ≥1 primary care GP or nurse consultation: £39 (Q1, Q3: £7, £39) and £16 (Q1, Q3: £16, £16), respectively (**Table 5**). These costs did not differ across the age, risk definition and immunocompromised status stratification for either the F2F or telephone consultations. The costs associated with treatment in the primary care setting were also analysed. Given the low prescribing associated with COVID-19 diagnoses in the primary care cohort, the associated medication costs were negligible, with the exception of costs among those who were immunocompromised (median cost: £21; Q1, Q3: £3, £566).

**Table 5:** Costs in the four weeks after index stratified by age, risk of severe COVID-19 and immunocompromised status at baseline among the primary care cohort.

| Costs |  | Age stratifications |  |  |  |  | Risk criteria |  |  | Immunocompromised status at baseline |  |
| --- | --- | --- | --- | --- | --- | --- | --- | --- | --- | --- | --- |
|  | All (n=1,693,263) | 18-49 (n=1,161,843) | 50-64 (n=379,528) | 65-74 (n=92,573) | 75-84 (n=40,481) | ≥85 (n=18,838) | McInnes Advisory Group (n=250,329) | PANO-RAMIC (n=691,593) | Green Book (n=212,556) | Immunocompromised (n=63,547) | Immunocompetent (n=1,629,716) |
| **Medication cost (£)** |  |  |  |  |  |  |  |  |  |  |  |
| Mean (SD) | 165 (465) | 242 (538) | 204 (505) | 134 (447) | 187 (530) | 8 (36) | 206 (518) | 159 (457) | 161 (448) | 486 (759) | 83 (307) |
| Median (Q1, Q3) | 2 (2, 4) | 3 (2, 256) | 2 (2, 24) | 2 (2, 3) | 3 (1, 10) | 3 (1, 3) | 3 (2, 21) | 2 (2, 4) | 3 (2, 9) | 21 (3, 566) | 2 (2, 3) |
| **Primary care consultations- F2F (£)** |  |  |  |  |  |  |  |  |  |  |  |
| Mean (SD) | 27 (22) | 26 (21) | 28 (22) | 27 (24) | 27 (25) | 27 (24) | 28 (24) | 28 (23) | 28 (25) | 28 (25) | 27 (22) |
| Median (Q1, Q3) | 39 (7, 39) | 39 (7, 39) | 39 (7, 39) | 39 (7, 39) | 39 (7, 39) | 39 (7, 39) | 39 (7, 39) | 39 (7, 39) | 39 (7, 39) | 39 (7, 39) | 39 (7, 397) |
| **Primary care consultations- Telephone (£)** |  |  |  |  |  |  |  |  |  |  |  |
| Mean (SD) | 18 (10) | 17 (9) | 18 (10) | 18 (10) | 19 (11) | 20 (13) | 18 (11) | 18 (10) | 19 (11) | 18 (10) | 18 (9) |
| Median (Q1, Q3) | 16 (16, 16) | 16 (16, 16) | 16 (16, 16) | 16 (16, 16) | 16 (16, 16) | 16 (16, 16) | 16 (16, 16) | 16 (16, 16) | 16 (16, 16) | 16 (16, 16) | 16 (16, 16) |

## DISCUSSION

To the best of our knowledge, this is the first study to quantify HCRU and related costs specific to the acute phase of COVID-19 in all adults (both standard and high-risk) within the primary and secondary care settings in England. Costs and HCRU were primarily driven by COVID-19 associated hospitalisations, particularly among older adults and those admitted to critical care, which imposed direct medical cost and resource use burden on the UK healthcare system.

Our findings on the overall LoS (6.0 days) were consistent with national estimates, indicating between August 2020 to March 2021 the median LoS ranged from 4-11 days.^35^ However, our data only covered early waves of the COVID-19 pandemic, where LoS fluctuated over time due to varying factors such as variant predominance, changes in COVID-19 testing guidance and COVID-19 vaccinations.^36^ For the patients admitted to critical care, we observed a median LoS of 8 days. A retrospective cohort study of patients admitted to ICU between March and May 2020, using COVID-19 Hospitalisation in England Surveillance System (CHESS) data found median LoS ranged from 10-12 days.^37^ This lower LoS observed in our study might partly be explained by refinements made to the treatment and management of COVID-19 patients over the course of the pandemic, starting with the publication of critical care guidance in March 2020.^38 39^ Further, we found critical care LoS was not monotonic with age and therefore its associated costs, which is also aligned with previous studies.^37 40^ It is possible that the LoS observed in patients aged ≥85 years was biased due to survivorship with a shorter apparent LoS due to an increased risk of death in the older ages. The proportion of patients admitted to critical care (14.8%) and requiring MV (6.0%) were also consistent with published English estimates over a longer data coverage period (10.6% and 5.6%, respectively).^41^

Our cost estimates associated with COVID-19 hospitalisation are similar to those used by the National Institute for Health and Care Excellence (NICE) Technology Appraisal assessing therapeutics for people with COVID-19, although the report restricted to severe COVID-19 patients and used non-comparable methodology.^42^.Data from other countries, for instance a retrospective study of hospitalised COVID-19 patients in the US from 1 April to 31 December 31 2020 using claims data, estimated the average cost per day for overall admissions and ICU admissions was $1,772 and $2,902, respectively,^43^ with evidence from Italy reporting the hospitalisation cost per day varying based on the complexity of care (low-complexity = €476; medium-complexity = €700; high-complexity = €1,402).^44^ However, the generalisability of these estimates to the UK population are limited given differences in populations, data coverage period, COVID-19 management strategies, and healthcare systems. Notably, due to variability within the relatively smaller immunocompromised and hospitalised, it is likely that our findings related to costs in this group differ to existing studies.^45-47^ However, as these patients are at an increased risk of severe COVID-19, they are likely to experience high HCRU and costs. Also, very minimal differences in HCRU were found across the different high-risk groups that were either prioritized for treatment (Highest risk group), eligible for the PANORAMIC clinical trial (PANORAMIC eligibility) or eligible for vaccination prioritization (Green book). All three subgroups incurred similar non-critical and critical care hospitalisation costs.

In the primary care setting, several major changes in the use of healthcare services occurred since the start of the pandemic, including: a reduction in health services (postponing non-urgent planned treatment and redeployment of NHS staff),^48^ an increased use of telemedicine resulting from a change in policy (F2F appointments only when clinically necessary),^49^ and a reluctance among patients to seek F2F care.^50-52^ These factors may explain the higher use of GP or nurse telephone consultation across both study cohorts, and overall limited use of GP or nurse consultations amongst the primary care cohort.

Our findings demonstrated that COVID-19 related hospitalisation continues to pose substantial pressure and cost to the healthcare system in England.^9^ This study reinforces the importance of continuing efforts with the UK COVID-19 vaccination program in reducing hospitalisation and severity of the disease.^53^ Policy makers and healthcare professionals should persist with encouraging high vaccination coverage, specifically among vulnerable groups and those at higher risk of hospitalisation with COVID-19.^54^

### Limitations of the study

Our study has several limitations. First, whilst CPRD covers 24% of the population in England,^55^ our previous published work found an underrepresentation of COVID-19 patients aged ≥65 years, particularly in the hospitalised cohort, and overrepresentation of patients living in specific English regions, such as London and the South East, for these two cohorts^19^ and thus our findings should be interpreted with caution. Second, due to data latency, HES APC data was only available up to March 2021 and emergency department as well as outpatient data were unavailable. For patients in the primary care cohort after April 2021, hospitalization status is unknown and therefore are unable to describe the HCRU and costs associated with more recent waves of the coronavirus pandemic. Cost estimates in the hospitalised immunocompromised patients should be interpreted with caution due to the smaller sample size. Lastly, our findings on the limited primary care prescriptions for the treatment of COVID-19 are expected given the first antiviral for COVID-19 in the UK was approved in November 2021,^56^ which occurred towards the end of the study period,^57^ the UK’s stringent access criteria when compared to other countries, and due to our definition of medication use (prescription on the same day as a COVID-19 diagnosis).

Future studies with a larger sample size are required to better quantify the economic burden of COVID-19 among immunocompromised patients. Studies should also explore the consequences of post-COVID conditions, to incorporate other aspects of HCRU e.g., readmissions, and the indirect costs associated with COVID-19 such as employment related sickness absence. Further, a better understanding of the health and social care needs of patients recovering from or who continue to experience symptoms of COVID-19 is required.

## CONCLUSION

The present retrospective cohort study quantified COVID-19 HCRU and associated costs in England. Although the burden of COVID-19 has reduced following the rollout of COVID-19 vaccines in the UK, we observed substantial economic burden due to COVID-19 on the NHS. Importantly, this study showed much of the burden during the study period was driven by COVID-19 related hospitalisations, and that older adults are associated with higher burden. Findings from this study can be used to inform the long-term strategy for resource allocation in the management of COVID-19.

## WHAT IS ALREADY KNOWN ON THIS TOPIC

- Since the start of the pandemic, COVID-19 has been associated ^∼^982,000 hospitalisations and ^∼^186,000 deaths in England
- Whilst the UK vaccination rollout has significantly reduced COVID-19 related morbidity and mortality, COVID-19 related hospitalisations and the associated impact remain high

## WHAT THIS STUDY ADDS

- During the acute phase of COVID-19 prior to omicron dominance, the majority of patients were managed within the primary care setting
- Hospitalisations, critical care admissions and increasing age (up to 65 years) were drivers of the higher healthcare resource use and related costs associated with COVID-19 in England
- Immunocompromised patients are likely to have high COVID-19 related HCRU and costs, however studies with larger sample size are needed to confirm this
- Very minimal differences in HCRU were found across the different high-risk groups that were either prioritized for treatment (McInnes Advisory Group), eligible for the PANORAMIC clinical trial (PANORAMIC) or eligible for vaccination (Green Book).

## Data Availability

Data may be obtained from a third party and are not publicly available. Anonymised patient data were accessed under study-specific approvals. Electronic health records are considered sensitive data in the UK by the Data Protection Act and cannot be shared. Access to the primary care data and linked datasets could be requested from the Clinical Practice Research Datalink https://www.cprd.com/research-applications.

## Acknowledgments

Editorial/medical writing support under the guidance of the authors was provided by Dr Gary Sidgwick PhD of Adelphi Real World, Bollington UK, in accordance with Good Publication Practice 2022 (GPP 2022) guidelines (https://www.ismpp.org/gpp-2022).

The authors gratefully acknowledge Tamuno Alfred, Darren Kailung Jeng and Chern Chuan Soo from Pfizer Inc. (New York, United States), Agnieszka Gajewska, Tomasz Mikołajczyk, Ewa Śleszyńska-Dopiera from Quanticate (Warsaw, Poland) and Olivia Massey from Adelphi Real World for statistical programming support.

## Copyright

The corresponding author has the right to grant on behalf of all authors and does grant on behalf of all authors, a worldwide licence to the Publishers and its licensees in perpetuity, in all forms, formats and media (whether known now or created in the future), to i) publish, reproduce, distribute, display and store the Contribution, ii) translate the Contribution into other languages, create adaptations, reprints, include within collections and create summaries, extracts and/or, abstracts of the Contribution, iii) create any other derivative work(s) based on the Contribution, iv) to exploit all subsidiary rights in the Contribution, v) the inclusion of electronic links from the Contribution to third party material where-ever it may be located; and, vi) licence any third party to do any or all of the above.

## Author contributions

The corresponding author, Jingyan Yang, attests that all listed authors meet authorship criteria and that no others meeting the criteria have been omitted.

All authors were involved in: 1) conception or design, or analysis and interpretation of data; 2) drafting and revising the article; 3) providing intellectual content of critical importance to the work described; and 4) final approval of the version to be published, and therefore meet the criteria for authorship in accordance with the International Committee of Medical Journal Editors (ICMJE) guidelines. In addition, all named authors take responsibility for the integrity of the work as a whole and have given their approval for this version to be published.

## Ethical approval

CPRD’s Research Data Governance (RDG) committee approved this study (CPRD study ID: 22_002062) in July 2022 prior to obtaining the data relevant to the project. This study complied with all applicable laws regarding subject privacy. As all patient-level data were fully anonymised, and no direct patient contact or primary collection of individual patient data occurred, patient consent was not required.

## Dissemination to participants and related patient and public communities

The study findings will be disseminated to the public through appropriate channels.

## Data sharing statement

### Data may be obtained from a third party and are not publicly available

Anonymised patient data were accessed under study-specific approvals. Electronic health records are considered sensitive data in the UK by the Data Protection Act and cannot be shared. Access to the primary care data and linked datasets could be requested from the Clinical Practice Research Datalink https://www.cprd.com/research-applications.

### Transparency declaration

The lead author affirms that this manuscript is an honest, accurate, and transparent account of the study being reported; that no important aspects of the study have been omitted; and that any discrepancies from the study as planned (and, if relevant, registered) have been explained.

### Prior publication

Data in this manuscript were accepted as three separate poster presentations at the International Society for Pharmacoeconomics and Outcomes Research (ISPOR) 2023, Boston, USA, in May 2023.

