## Supplementary Materials for "Healthcare resource utilisation and costs of hospitalisation and primary care among adults with COVID-19 in England: a population-based cohort study"

**Supplementary eFigure 1** Study design schematic

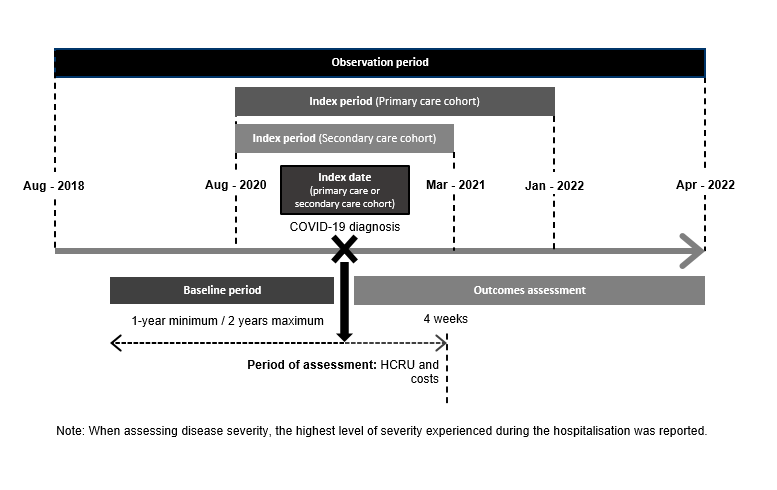

**Supplementary eTable 1** COVID-19-related medications

**Generic name**

Nirmatrelvir/ritonavir

Sotrovimab

Molnupiravir

Casirivimab/imdevimab

Remdesivir

Dexamethasone

Tocilizumab

Sarilumab

Baricitinib

**Supplementary eTable 2** Primary care HCRU stratified by age, risk of severe COVID and immunocompromised status at baseline among the hospitalised cohort

| **HCRU** |  | **Age stratifications** | | | | | **Risk criteria** | | | **Immunocompromised status at baseline** | |
| --- | --- | --- | --- | --- | --- | --- | --- | --- | --- | --- | --- |
|  | **All**  **(n=13,105)** | **18-49 (n=3,127)** | **50-64 (n=4,844)** | **65-74 (n=2,386)** | **75-84 (n=1,690)** | **≥85 (n=1,058)** | **McInnes Advisory Group (n=4,323)** | **PANORAMIC (n=11,011)** | **Green Book**  **(n=** **5,341)** | **IC**  **(n=181)** | **Non-IC (n=12,924)** |
| **Any COVID-19 medication use:** n, (%) | 29  (0.2%) | 6 (0.2%) | 9 (0.2%) | 5 (0.2%) | 5 (0.3%) | <5 | 14  (0.3%) | 27  (0.3%) | 15  (0.3%) | <5 | 28  (0.2%) |
| **Primary care consultations- F2F:** number with ≥ 1 visit (%) | 2,489 (19.0%) | 559 (17.9%) | 921 (19.0%) | 425 (17.8%) | 354 (21.0%) | 230 (21.7%) | 900 (20.8%) | 2,124 (19.3%) | 1,093 (20.5%) | 31  (17.1%) | 2,458 (19.0%) |
| **Primary care consultations- Telephone:** number with ≥ 1 call (%) | 5,077 (38.7%) | 1,178 (37.7%) | 1,840 (38.0%) | 949 (39.8%) | 701 (41.5%) | 409 (38.7%) | 1,815 (42.0%) | 4,341 (39.4%) | 2,306 (43.2%) | 87  (48.1%) | 4,990 (38.6%) |

**IC: immunocompromised; Non-IC: non-immunocompromised also known as immunocompetent.**

**Supplementary eTable 3** Primary care costs stratified by age, risk of severe COVID and immunocompromised status at baseline among the hospitalised cohort

|  |  | **Age stratifications** | | | | | **Risk criteria** | | | **Immunocompromised status at baseline** | |
| --- | --- | --- | --- | --- | --- | --- | --- | --- | --- | --- | --- |
|  | **All**  **(n=** **13,105)** | **18-49**  **(n=** **3,127)** | **50-64**  **(n=** **4,844)** | **65-74**  **(n= 2,386)** | **75-84**  **(n=** **1,690)** | **≥85 (n=1,058)** | **McInnes Advisory Group**  **(n=** **4,323)** | **PANORAMIC (n=** **11,011)** | **Green Book**  **(n=** **5,341)** | **IC**  **(n=181)** | **Non-IC (n=12,924)** |
| **Medication cost (£)** |  |  |  |  |  |  |  |  |  |  |  |
| Mean (SD) | 4.8 (15.4) | 1.8 (1.1) | 1.9 (1.5) | 2.6 (1.4) | 2.2 (1.3) | 22.2 (41.6) | 8.2 (22.0) | 5.1 (15.9) | 7.7 (21.3) | 0.6 (-) | 5.0 (15.7) |
| Median  (Q1, Q3) | 1.8 (1.2, 2.5) | 1.5 (0.9, 2.5) | 1.3 (1.2, 1.8) | 1.8 (1.8, 2.5) | 2.5 (1.3, 2.8) | 1.9 (0.9, 43.6) | 2.5 (1.3, 3.6) | 1.8 (1.2, 2.5) | 2.5 (1.3, 3.6) | 0.6 (0.6, 0.6) | 1.8 (1.2, 2.5) |
| **Primary care consultations- F2F (£)** |  |  |  |  |  |  |  |  |  |  |  |
| Mean (SD) | 30.5 (26.5) | 31.4 (30.3) | 31.9 (24.7) | 30.5 (26.3) | 28.7 (28.1) | 25.5 (20.1) | 30.0 (24.6) | 30.4 (25.8) | 30.0 (25.6) | 35.6 (27.1) | 30.4 (26.5) |
| Median  (Q1, Q3) | 39.2  (6.8, 39.2) | 39.2  (6.8, 39.2) | 39.2  (6.8, 39.2) | 39.2  (6.8, 39.2) | 39.2  (6.8, 39.2) | 13.5  (6.8, 39.2) | 39.2  (6.8, 39.2) | 39.2  (6.8, 39.2) | 39.2  (6.8, 39.2) | 39.2  (13.5, 39.2) | 39.2  (6.8, 39.2) |
| **Primary care consultations- Telephone (£)** |  |  |  |  |  |  |  |  |  |  |  |
| Mean (SD) | 22.5 (13.8) | 23.2 (14.7) | 22.7 (13.6) | 22.2 (14.0) | 21.6 (13.1) | 21.4 (12.5) | 23.0 (14.2) | 22.4 (13.6) | 22.8 (13.9) | 23.0 (15.5) | 22.5 (13.8) |
| Median  (Q1, Q3) | 15.5  (15.5, 31.0) | 15.5  (15.5, 31.0) | 15.5  (15.5, 31.0) | 15.5  (15.5, 31.0) | 15.5  (15.5, 31.0) | 15.5  (15.5, 31.0) | 15.5  (15.5, 31.0) | 15.5  (15.5, 31.0) | 15.5  (15.5, 31.0) | 15.5  (15.5, 31.0) | 15.5  (15.5, 31.0) |

**IC: immunocompromised; Non-IC: non-immunocompromised also known as immunocompetent.**
